# Yield of Long-Read Genome Sequencing for Rare Disease Diagnosis in Short-Read Genome Negative Cases

**DOI:** 10.64898/2026.09.09.26362331

**Authors:** Georgia Pitsava, Krista Bluske, Rebekah Barrick, Ivan De Dios, Cathy Duong, Kirsten Blanco, Sami Belhadj, Nida Karra, Sarah L. Stenton, Arthur Garcia, Erica Smith, Miguel Almalvez, Arthur Ko, John Harting, Stuti Joshi, Kenny Chen, Charles Hadley King, Jonathan LoTempio, Jessica Albert, Kinga M. Bujakowska, Leandros Boukas, UCI-GREGoR Consortium, Rachid Karam, Seth I. Berger, Emmanuèle C. Délot, Changrui Xiao, Eric Vilain

**Author notes:** Corresponding author Eric Vilain, MD, PhD, Vice Dean, Clinical Research, School of Medicine Associate Vice Chancellor, Scientific Affairs, Health Affairs Director, Institute for Clinical and Translational Science Professor, Department of Pediatrics, University of California, Irvine, 1003 Health Sciences Road Suite 308, Irvine, CA 92617.

## Abstract

**Background:** A growing body of work has highlighted advantages of long-read genome sequencing (LR-GS) over short-read genome sequencing (SR-GS) with regards to the detection and interpretation of pathogenic variants. However, the incremental diagnostic yield of LR-GS over SR-GS for patients with suspected Mendelian conditions has not been systematically characterized.

**Methods:** We performed LR-GS on 144 cases that previously had undergone SR-GS through the Pediatric Mendelian Genomics Research Center at the University of California, Irvine (PMGRC-UCI, a founding center of the GREGoR Consortium), of which 107 remained undiagnosed after SR-GS.

**Results:** We identified new molecular diagnoses in 13 cases, five of which could not have been detected with SR-GS, corresponding to an incremental diagnostic yield of 4.7%. These five diagnoses leveraged distinct capabilities of LR-GS: the detection of variants in short-read “dark” regions, the detection of structural variants and tandem repeat expansions, and the identification of *de novo* variants using only one biological parent. In two of these five cases, confirmation of the diagnosis also relied critically on identifying aberrant transcript production via RNA-seq, while in one other case LR-GS-based CpG methylation profiling further supported the diagnosis through detection of abnormal methylation at episignature-associated CpGs.

**Conclusions:** In this paired comparison of LR-GS and SR-GS, LR-GS provided an incremental diagnostic yield of 4.7% among cases that remained undiagnosed after SR-GS. Together, our findings demonstrate that LR-GS can resolve a meaningful subset of cases beyond the reach of SR-GS while simultaneously expanding the range of genomic and epigenomic mechanisms accessible to a single sequencing assay.

## Background

Long-read genome sequencing (LR-GS) is emerging as a powerful advance over short-read genome sequencing (SR-GS) for the comprehensive characterization of human genetic variation. Despite the widespread adoption of SR-GS-based approaches, including clinical exome and genome sequencing, the diagnostic yield for rare genetic disorders is frequently reported as remaining below 50%, underscoring a substantial remainder of unsolved cases (Wojcik et al. 2023; Bruels et al. 2022; Negi et al. 2025), although exact estimates are hard to come by as they would require the systematic analysis of a naïve cohort. This diagnostic gap reflects both incomplete biological understanding, such as incomplete knowledge of gene-phenotype relationships, the impact of variants in non-coding regions of the genome, and fundamental technical limitations of SR-GS particularly in repetitive, homologous or low complexity regions and in the detection of complex structural variation.

Over the past few years, several benefits of LR-GS have become apparent. First, by generating reads spanning from ∼ 10kb to over 1 Mb, LR-GS enables more accurate mapping across repetitive loci and increases our ability to detect structural variants, repeat expansions, and variants in highly homologous regions (Mahmoud et al. 2024; Logsdon et al. 2020; LoTempio et al. 2023; Dawood et al. 2025). Second, it allows the detection of not just genome sequence variation, but also variation in epigenetic modifications such as DNA methylation (Ni et al. 2023; Fu et al. 2025; Mortazavi et al. 2026). Third, it permits direct, read-backed phasing, making it possible to characterize genetic variants as well as chromatin states at the haplotype level (Miller et al. 2021; Cohen et al. 2022; Stergachis et al. 2020; Vollger et al. 2025; Steyaert et al. 2025). The phasing capability of LR-GS has also recently been shown to enable the identification of *de novo* variants without requiring sequencing of both biological parents, highlighting a common situation where LR-GS can improve not only the detection, but also the interpretation of variants (Boukas et al. 2026). Together, these benefits position LR-GS as a promising strategy for identifying causative variants in a greater proportion of patients with rare conditions than SR-GS.

Several studies have highlighted cases solved after LR-GS uncovered pathogenic variants missed by SR-exome or genome sequencing (Bruels et al. 2022; Sheth et al. 2026; Liang et al. 2026; Steyaert et al. 2025; de Bitter et al. 2026; Mortazavi et al. 2026). However, there are limited data on the incremental diagnostic yield of LR-GS when it is applied in an unbiased fashion in a rare disease cohort that has previously undergone systematic evaluation via SR-GS. Here, we report LR–GS results from 144 families (107 unsolved, total 371 individuals) for whom we also had access to prior SR-GS results. We assess the added diagnostic yield from LR-GS over SR-GS, highlighting representative cases and types of variants.

## Methods

### Participants

Participants were recruited into the PMGRC (Pediatric Mendelian Genomics Research Center) research project as part of the GREGoR (Genomics Research to Elucidate the Genetics of Rare diseases)) Consortium. Inclusion criteria included probands with a suspected Mendelian condition who had undergone prior standard-of-care clinical testing, including gene panels, chromosomal microarray, and/or exome sequencing, and had non-diagnostic results. Testing on additional informative family members was performed when available. All work was done under Institutional Review Board (IRB) protocols Pro00015852 at Children’s National Hospital and #4469 at the University of California, Irvine. Informed consent was obtained from all participants. SR-GS and LR-GS were performed sequentially. Individuals were enrolled over a three-year period, from July 2021 to March 2024. Buccal swabs and/or peripheral blood samples were collected from probands and relevant available family members. Phenotypic data were obtained through manual review of electronic medical records or provided by collaborating clinicians and were standardized using Human Phenotype Ontology (HPO) terms. SR-GS methods in this cohort were previously published (Pitsava et al. 2025). Data were held and shared internally via a custom-built dashboard, designed to support phenotype collection, sample tracking, sequencing metadata standardized to the GREGoR data model for rare disease genomics research, links to datasets repositories, and analysis reports (King et al. 2026).

### Long-read Genome Sequencing

High-molecular-weight (HMW) genomic DNA (gDNA) was isolated using the Nanobind HT CBB kit for blood specimens (PacBio, Menlo Park, CA, USA), and the Quick-DNA HMW MagBead Kit for buccal/saliva specimens (Zymo Research, Irvine, CA, USA). Automated pipette shearing of 1-2 µg of gDNA was performed on Tecan Fluent 1080 instruments (Tecan, Männedorf, Switzerland) to obtain a fragment size distribution peak between 12-22 kb. Library preparation was performed using the HiFi Prep 96 kit (PacBio, Menlo Park, CA, USA) following the manufacturer-recommended protocol, followed by gel-based size selection on a LightBench instrument (YourGene Health, Manchester, United Kingdom). Sequencing was performed on PacBio Revio instruments by Ambry Genetics, targeting an average sequencing depth of ≥20x and a median read quality of ≥Q30.

### Data preparation for analysis

The workflow developed for this study is illustrated in Figure 1. Variant prioritization and case analysis were performed on the GeneYX platform (Geneyx Inc., Wilmington, DE), which allowed centralization of the findings of multiple analysts using standard operating procedures. Raw data (unaligned HiFi reads) were processed using the PacBio HiFi-human-GS-WDL pipeline (https://github.com/PacificBiosciences/HiFi-human-GS-WDL), using version 1.0 for earlier samples, and version 2.0/3.0 for the most recent samples.

**Figure 1.**
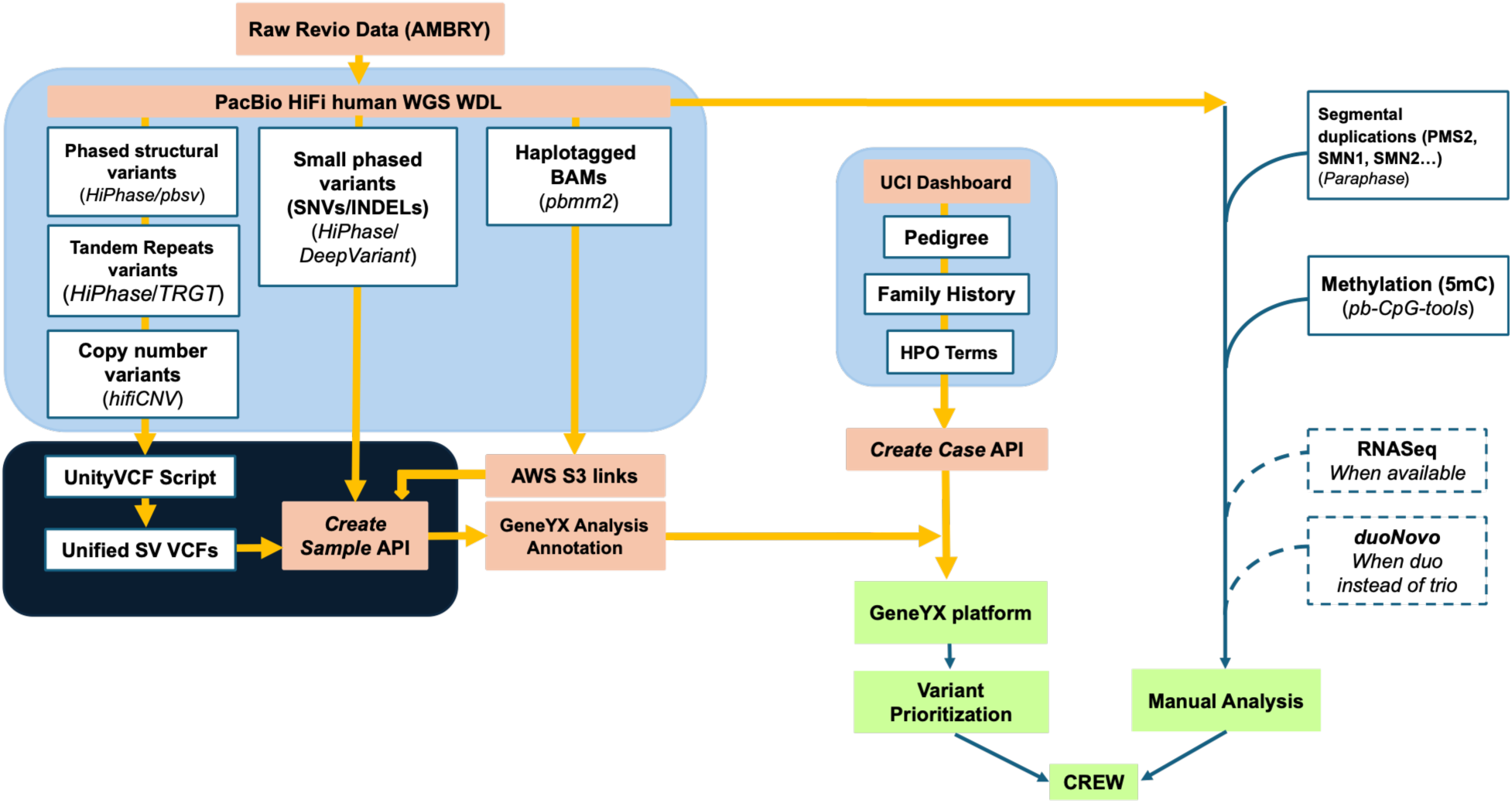
Workflow for long-read sequencing and analysis. HiFi sequencing was performed on a Revio instrument at Ambry Genetics. Variant prioritization was performed on the GeneYX platform. Processing of raw data through the PacBio WDL yielded VCFs from the various tools used to detect SNV/indels, structural variants, copy-number variants, and tandem repeats. For analysis we additionally used a dashboard to document family history and phenotype according to the data model developed by the GREGoR Consortium specifically for rare disease research. Other data available in the HiFi sequencing dataset, such as methylation calls or output of the *Paraphase* tool are analyzed manually. When available, RNASeq datasets were used to evaluate the impact of variants on mRNA splicing and stability.

For each HiFi sequence, the VCF outputs of *DeepVariant* (single nucleotide variants, SNVs and Indels), *pbsv* (structural variants, SVs), *hifiCNV* (copy-number variants, CNVs), and *TRGT* (tandem repeats) tools were downloaded from our AWS S3 bucket storage. The *pbsv, hifiCNV,* and *TRGT* VCFs were merged into a unified SV VCF using a Geneyx-provided python script.

For each participant, all files, together with the list of HPO terms, were then uploaded to the analytical platform via web APIs referencing a subject ID. Cases were subsequently created to associate family members with the proband.

### Variant filtering

“Smart Filtering” v6.2 was applied to remove common and low-quality variants from SNV calls. Smart Filtering parameters included filtering variants using an hg38 bed file generated from RefSeq hg38 transcript annotations (UCSC ncbiRefSeq.txt.gz, downloaded June 2025). All NM_ and a curated subset of NR_ transcripts from coding genes were included and transcript intervals were merged per gene across chromosomes 1–22, X, Y, and M (mitochondrial genome). The resulting gene-level regions were padded by ±5 kb to capture nearby regulatory elements and support gene-centric variant filtering. GnomAD v4.0 filtering by allele counts worked as follows: homozygous alleles present in 50 or more individuals, heterozygous alleles present in 6,000 or more individuals, and hemizygous alleles present in 10 or more individuals.

### Variant annotation

After variant filtering, SNV/Indel annotation was performed using CADD 1.7, SpliceAI 1.3, and SnpEff (Cingolani 2022; Rentzsch et al. 2019; de Sainte Agathe et al. 2023). Non-coding variants with SpliceAI scores less than 0.05 were also excluded. Structural variants (SVs) from pbsv (Yuan and Jia 2024) were annotated with needLR (Gustafson et al. 2026). For tandem repeats, aligned genome bam files were processed through LR tandem repeat genotyping (*TRGT*) version 3.0.0 with reference loci adotto_strchive_20250626. Known disease-causing loci (72) were selected from those published in STRchive with the pathogenic ranges extracted from the STRchive json file (L. Hiatt et al. 2025). Variants with pathogenic repeat units numbering within the pathogenic range were flagged for manual review by a team of variant analysts.

### Methylation analysis

The approach we have previously described in (Negi et al. 2025) was followed. Methylation levels were assessed at each of the CpGs comprising published rare disease episignatures. Samples were grouped in batches based on SMRTcell type and Revio Jasmine methylation caller version to minimize technical batch effects. A concordance score was generated for each individual sample through taking the cosine similarity of the methylation calls with published beta-values of the episignature for the condition of interest. These scores were then z-transformed to assess each sample for being an outlier with regards to their methylation status at these CpGs compared to all other individuals in our cohort sequenced on the same chemistry, with the same library preparation, and called using the same version of Jasmine. A sample was considered an outlier if its score was greater than 2.5 standard deviations above the mean for the batch.

### Case Level Analysis

A team of variant analysts utilized custom filters and genotype- and phenotype-driven prioritization to evaluate the clinical relevance of variants (Austin-Tse et al. 2022). Interpretation of identified variants followed ACMG/AMP criteria and ClinGen recommendations (Richards et al. 2015; Riggs et al. 2020). Other data available in the HiFi sequencing dataset, such as methylation calls or output of the *Paraphase* tool (to disambiguate segmental duplications) (X. Chen et al. 2025), were analyzed manually as needed. When available, RNASeq datasets were used to assess the impact of variants on mRNA splicing. Prioritized variants were discussed during a weekly meeting of a multidisciplinary team.

### Definitions

Following standardized definitions from the GREGoR consortium (Heavner et al. 2026), cases were classified as “solved” when pathogenic/likely pathogenic variant(s) were identified in a gene associated with a condition consistent with the proband’s presentation, and when the number and phase of variants were concordant with the expected inheritance pattern (e.g., a single pathogenic or likely pathogenic variant for autosomal dominant conditions, or biallelic pathogenic or likely pathogenic variants for autosomal recessive conditions). Cases were considered “probably solved” when one or more variants of uncertain significance (VUS) in characterized genes were identified and there was strong phenotypic concordance, or when a pathogenic or likely pathogenic variant was identified but the phenotype was consistent yet not specific. Finally, cases were considered “partially solved” when pathogenic or likely pathogenic variants consistent with the inheritance pattern were detected but explained only a subset of the proband’s phenotype.

### Mini-gene splicing assay

When a candidate variant was found in a gene not expressed in blood or no RNA sequencing dataset was available, we deployed a mini-gene splicing assay to test the potential effect on splicing (Pitsava et al. 2025). This was used here to examine the impact of NM_005032.7: c.368-27A>C in *PLS3* found in proband PMGRC-590-590-0 (Supp. Fig. 1). Reference and variant (NM_005032.7(*PLS3*): c.368-27A>C) gene fragments containing 1000 bp of *PLS3* exons 4 and 5 (ENST00000355899.8) and surrounding intron sequence were inserted into mini-gene split GFP constructs. Following transfection, total RNA was extracted, reverse-transcribed, and amplified to visualize splicing events on electrophoresis (detailed methods in Supp. Fig. 1) (Scott et al. 2022; Chong et al. 2019).

### duoNovo

When only one biological parent sample was available, the *duoNovo* tool was used to classify candidate variants, where the proband is heterozygous and the sequenced parent is homozygous for the reference allele. *duoNovo* uses surrounding haplotype sequence similarity to infer whether each candidate variant is present on the haplotype transmitted from the sequenced parent, thus allowing inference of whether it is *de novo* or not. Default parameters were used for genome-scale analysis as described in (Boukas et al. 2026). Specifically, the R package was run using the corresponding command-line interface (https://github.com/sbergercnmc/duonovo).

## Results

In total, 144 families (371 participants in total, 144 probands) underwent both SR-GS and LR-GS. Sex, race, and phenotype distribution of probands who underwent LR-GS is shown in Figure 2. Males exceeded females among probands by about 13% (**Fig. 2A**). The cohort was ethnically diverse, with the majority reported being either white (∼60%) or of more than on race (∼22%) (**Fig. 2B**). Phenotypes were also very diverse (**Fig. 2C**), with the most common phenotype among probands being developmental delay, likely explaining the male bias composition of the cohort. Family structures included 20 proband-only families, 36 duos or duos with another (non-parent) family member (designated as duo+), 87 trios or trios with another family member (trio+), and one family consisting only of 2 siblings. As positive control, we included 37 cases that previously had a positive finding by SR-GS (28 were solved, nine harbored a candidate variant; Supp. Table 1; (Pitsava et al. 2025)) and confirmed that LR-GS was able to identify the same diagnostic finding following appropriate manual curation (Supp. Table 1).

**Figure 2.**
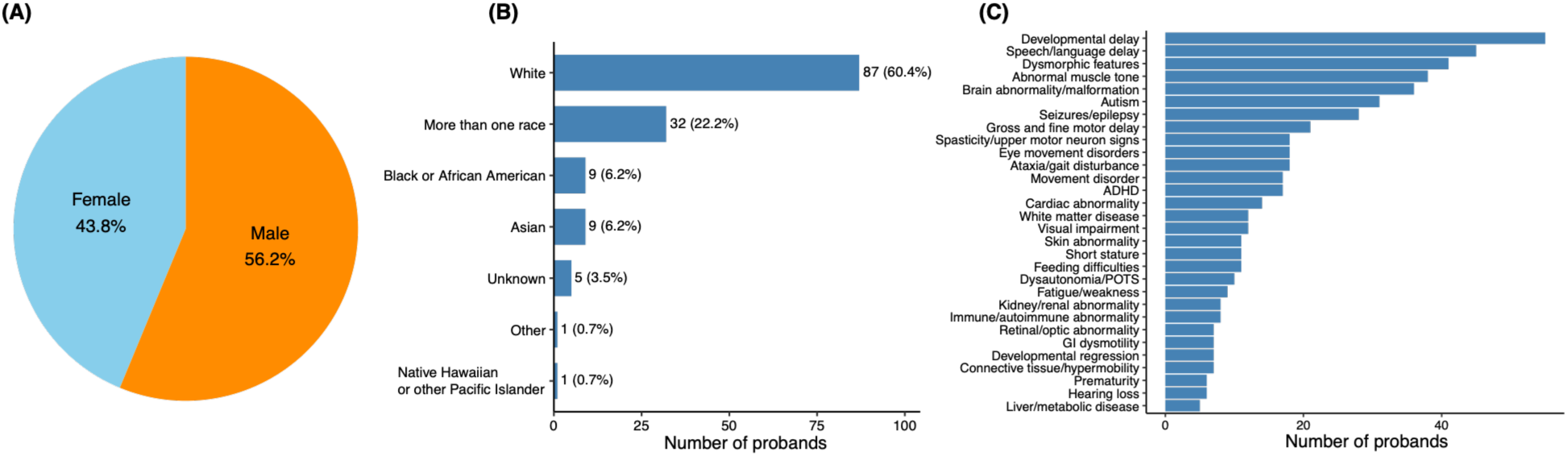
Characteristics of the cohort. **(A), (B)** Sex and race distribution of probands who underwent long read genome sequencing. **(C)** Number of probands with different phenotypes in our cohort.

We found a diagnosis in 13 (including three partially solved cases, unambiguously explaining part, but not all the phenotype) out of 107 unsolved cases (12.1%, **Fig. 3A**). Upon review, eight cases received a diagnosis that could have been discovered with reanalysis of prior SR data (indicated with a double asterisk in the first column of Table 1). However, five cases received a diagnosis that could not have been detected with SR-GS, consistent with an incremental diagnostic yield of LR-GS over SR-GS of 4.7%. We describe these five cases below.

**Figure 3.**
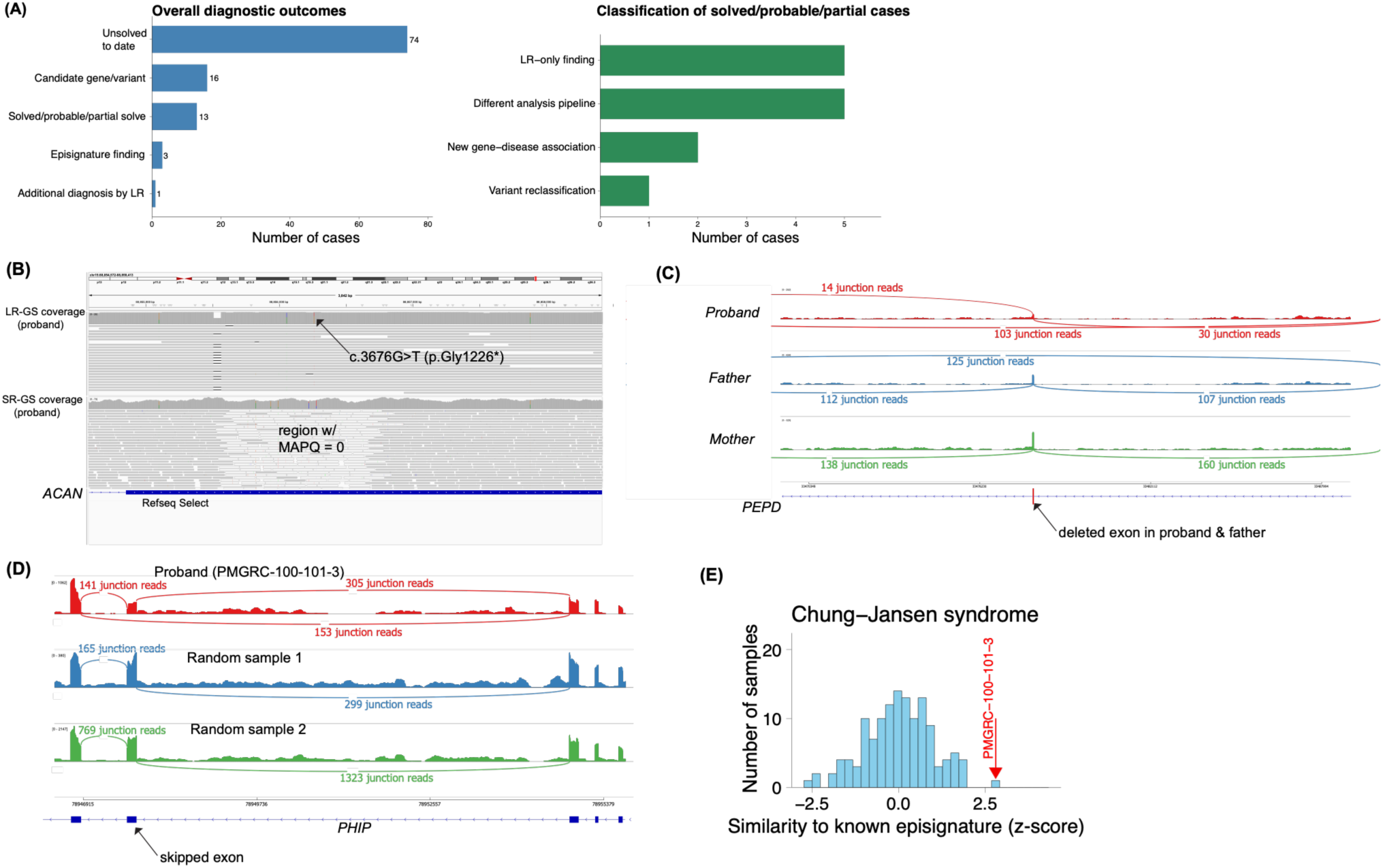
LR-GS delivered diagnoses that could not have been found with SR-GS in 4.7% of cases. **(A)** Breakdown of the cohort according to solve status (left) and reason why our study identified a new diagnosis. **(B)** IGV snapshot showing HiFi aligned reads in the region containing the pathogenic variant in *ACAN* revealed by LR-GS in participant PMGRC-1286-1286-0 (top panel). The bottom panel shows the same region on the proband’s SR-GS with a large portion of exon 12 unmappable with short reads. **(C)** IGV sashimi plot and splice junction plot depicting junctions of *PEPD* exons 6, 7, and 8 in the proband (PMGRC-645-645-0, red), the father (PMGRC-646-645-1, blue), and the mother (PMGRC-644-645-2, green) from RNA-seq. The IGV splice junction plot is normalized to a read depth of 350X for the proband, father, and mother. The proband and father show the exon 7 skipping junction reads (103 reads joining exon 6 to exon 8 in the proband, 125 such junction reads in the father). Junction reads joining exon 6 to exon 8 are entirely absent from the mother. **(D)** Sashimi plot for PMGRC-100-101-3 (shown in red at the top) and for two random samples (shown in blue and green below) sequenced in the sample batch. The proband was found to harbor a *de novo* intronic variant in *PHIP*. In this sample, 153 junction reads at the acceptor and donor sites supported exon skipping, which was not observed in the two random samples (RNA-seq from the mother and the sibling in this family were not available). **(E)** Similarity z-score for PMGRC-100-101-3 (red arrow), against the distribution of similarity z-scores for the rest of the cohort (n=130) indicate that the sample is an outlier for CpG methylation levels at the 230 positions included in the Chung-Jansen Syndrome episignature

**Table 1.** Solved/probable solved/partially solved cases by long-read genome sequencing.

| Participant ID<br>(Case number in<br>this report) | Gene<br>(MIM ID;<br>transcript<br>ID) | Variant (c.<br>or n.) | Variant<br>(p.) | Variant<br>(g.) | Inheritance | Patient Phenotype | Associated<br>Disorder<br>(MIM,<br>ORPHA) | ACMG<br>Classification |
| --- | --- | --- | --- | --- | --- | --- | --- | --- |
| PMGRC-100-101-3<br>(Case 3) | *PHIP*<br>(#612870,<br>NM_01793<br>4.7) | c.4206+3A>G | p.? | NC_000006.12:g.78947620T>C | De novo | Developmental delay, speech delay | Chung-Jansen syndrome (#617991) | LP |
| PMGRC-423-423-0** | *FGF8*<br>(#600483,<br>NM_033163.5) | c.290_337dup | p.(Phe112_Ala113insValAlaAsnLysArgIleAsnAlaMetAlaGluAspGlyAspProPhe) | NC_000010.11:g.101774732_101774779dup | Paternally inherited | Holoprosencephaly, nonverbal, non-ambulatory | Holoprosencephaly (ORPHA:220386) | LP |
| PMGRC-431-431-0** | *FGF8*<br>(#600483,<br>NM_033163.5) | c.356C>T | p.Thr119Met | NC_000010.11:g.101771551G>A | Paternally inherited | Holoprosencephaly, cerebral palsy, cognitive delay | Holoprosencephaly (ORPHA:220386) | LP |
| PMGRC-559-559-0** | *GIGYF1*<br>(#612064,<br>NM_001375765.1) | c.332del | p.Leu111fs | NC_000007.14:g.100687546del | De novo | Chiari malformation, autism, global developmental delay, seizures, dystonia, ataxia, cafe-au-laits | GIGYF1-related syndrome | P |
| PMGRC-562-562-0** | *TSC2*<br>(#191092,<br>NM_000548.5) | c.2648A>G | p.(Gln883Arg) | NC_000016.10:g.2079395A>G | Unknown | Seizures, temporal dysplasia, multiple fractures, neurocognitive delay, autism, motor skills regression, obesity | Focal cortical dysplasia, type II (#607341) | VUS |
| PMGRC-572-572-0** | *RPL31*<br>(#617415,<br>NM_000993.5) | c.-1+3_-1+6del | p.? | NC_000002.12:g.101002318_101002321del | Unknown | Diamond Blackfan anemia, thumb abnormalities, radial hypoplasia |  | LP |
| PMGRC-590-590-0** | *PLS3*<br>(#300131,<br>NM_005032.7) | c.368-27A>C | p.? | NC_000023.11:g.115629808A>C | Maternally inherited | Decreased bone density, juvenile osteoporosis, recurrent fractures, high pain tolerance | Bone mineral density QTL18, osteoporosis (#300131) | LP |
| PMGRC-645-645-0<br>(Case 2) | PEPD<br>(#613230,<br>NM_000285.4) | c.769G>T | p.Gly257* | NC_000019.10:g.33411721C>A | Maternally inherited | Developmental delay, splenomegaly, eczema, intractable diarrhea, vomiting, recurrent skin abscesses, autoimmune hepatitis, dysmorphic features | Prolidase deficiency (#170100) | P |
|  |  | DEL at Chr19:33471512-33484890 | p.? |  | Paternally inherited |  |  |  |
| PMGRC-730-730-0** | RNU2-2<br>(#621238,<br>NC_000011.10) | n.4G>A | NA | NC_000011.10:g.62841806C>T | Unknown | Global developmental delay, macrocephaly, seizures, dysmorphic features | Developmental and epileptic encephalopathy 119 (#621304) | P |
| PMGRC-760-760-0** | PPP2R5D<br>(#601646,<br>NM_006245.4) | c.589G>C | p.Glu197Gln | NC_000006.12:g.43007271G>A | Paternally inherited | Global developmental delay, epilepsy, nonverbal, motor delay, hypotonia, self-injurious behavior, MRI: bilateral white matter injury and volume loss | Houge-Janssens syndrome 1 (#616355) | LP |
| PMGRC-1067-1067-0<br>(Case 5) | BEAN1<br>(#612051,<br>NM_001178020.3) | 615 TAGAA repeats |  |  | Paternally inherited | Increased fatigue, exercise intolerance, muscle weakness, malignant hyperthermia, unstable gait, decreased complex III and IV, chronic pain | Spinocerebellar ataxia 31 (#117210) | P |
| PMGRC-1284-1284-0<br>(Case 4) | CNBP<br>(#116955,<br>NM_003418.5) | 460 CAGG repeats |  |  | Nonmaternal | Cognitive fatigue, memory impairment, impaired executive functioning, paresthesia, anxiety, EDS, aortic root dilation, pancreatic insufficiency | Myotonic dystrophy 2 (#602668) | P |
| PMGRC-1286-1286-0<br>(Case 1) | ACAN<br>(#155760,<br>NM_001369268.1) | c.3676G>T | p.Gly1226* | NC_000015.10:g.88856261G>T | Paternally inherited | Osteochondritis dissecans lesions, short stature, ADHD | Short stature and advanced bone age, with or without early-onset osteoarthritis and/or<br>osteocondritis<br>dissecans<br>(#165800) | P |

*ADHD* attention-deficit/hyperactivity disorder, *LP* Likely pathogenic, *NA* not applicable, *P* pathogenic, *VUS* variant of uncertain significance, **\*\***indicates cases that would likely have been discoverable upon reanalysis of the SR-GS data.

### Identification of a pathogenic variant within a VNTR where SR-GS reads align with zero mapping quality

**Case 1 (PMGRC-1286-1286-0):** a young female with multiple osteochondritis dissecans lesions, short stature and ADHD. Family history was notable for osteochondritis and short stature in one parent, as well as severe and early-onset osteochondritis in the grandparent, suggesting an autosomal dominant pattern of inheritance. Previous genetic testing, including chromosomal microarray, exome sequencing and SR-GS, was negative. LR-GS identified a paternally inherited nonsense variant (NM_001369268.1:c.3676G>T (p.Gly1226*)) in *ACAN* **(Fig. 3B)**, a gene where loss-of-function variants are known to cause an autosomal dominant condition termed short stature with advanced bone age, with or without early-onset osteoarthritis and/or osteochondritis dissecans (SSOAOD; MIM#165800; (Stattin et al. 2010)). This variant was interpreted as pathogenic by ACMG/AMP criteria and deemed diagnostic. Further interrogation of the genomic region harboring this variant revealed that it was not detected by SR-GS because it fell within a 1-kb “dead zone” in exon 12, where SR-GS reads align with zero mapping quality due to a variable number tandem repeat of 57 bp typically repeated 13-44 times in the general population; (Roughley et al. 2006; Doege et al. 1997; Nakamura et al. 2025).

### Identification of a pathogenic deletion missed by SR-GS

**Case 2 (PMGRC-645-645-0)**: a young male with short stature, autoimmune enterocolitis and hepatitis leading to acute liver failure and subsequent transplant, splenomegaly, developmental delay, recurrent skin abscesses and scrotal hidradenitis suppurativa, and dysmorphic features. Prior genetic testing, including a primary immunodeficiency panel (containing 429 genes) and exome sequencing, was non-diagnostic. SR-GS identified a maternally inherited heterozygous stop-gain variant (NM_000285.4:c.769G>T (p.Gly257*)) in *PEPD,* previously classified as pathogenic in ClinVar (ID:2580569) and associated with prolidase deficiency (MIM#170100). While the patient’s phenotype was consistent with prolidase deficiency, this is an autosomal recessive disorder, and a diagnosis could not be established in the absence of an additional pathogenic variant in *trans*. LR-GS subsequently revealed a paternally inherited 13.4 kb (Chr19:33471512-33484890) deletion encompassing the in-frame exon 7 of *PEPD*. This deletion was missed by the SR structural variant callers (Manta, Delly, Lumpy, DRAGEN, Canvas, and Rainbow) and was only detectable in retrospect by manual inspection and supported by low-confidence calls. RNA-sequencing showed 50% expression of *PEPD* in the mother (consistent with the stop-gain variant causing nonsense-mediated decay), exon 7 skipping in the father, and the combination of those alterations in the proband (**Fig. 3C**), as evidenced by very low expression of exon 7. Notably, deletion of exon 7 is a known pathogenic mechanism for prolidase deficiency (Ledoux et al. 1994). Urine amino acid analysis using a Biochrom amino acid analyzer demonstrated imidodipeptiduria, biochemically confirming the diagnosis (Supp. Fig. 2A). The patient’s similarly affected sibling was also biochemically confirmed to have prolidase deficiency (Supp. Fig. 2B).

### LR-GS enabled both identification and validation of a de novo variant from a family with only one biological parent sequenced

#### Case 3 (PMGRC-100-101-3)

Two young male siblings were referred to the study: PMGRC-101-101-0 with hemihypertrophy and club foot and PMGRC-100-101-3 with developmental delay and dysmorphic features.

Because one of the parents had not provided a sample for research purposes, it was not possible to identify *de novo* variants in this family. However, cohort-wide application of *duoNovo*, a method we recently developed to leverage long reads to infer variants’ inheritance status even when one of the biological parents is not available for sequencing, had yielded a candidate *de novo* variant in the older sibling (Boukas et al. 2026). This was an intronic variant (NM_017934.7:c.4206+3A>G) in *PHIP*, which had arisen *de novo* on the maternal (sequenced) haplotype. Haploinsufficiency of *PHIP* is associated with the neurodevelopmental disorder Chung-Jansen syndrome (MIM#617991), which includes developmental delays, behavioral manifestations, and characteristic facial dysmorphism (Webster et al. 2016; Jansen et al. 2018). The variant had not been closely examined previously on SR-GS or LR-GS because its *de novo* status was unknown. After *duoNovo* classified it as *de novo*, we sought to assess the variant’s effect through orthogonal methods. It was predicted to affect splicing, albeit with somewhat inconsistent scores. *SpliceAI* predicted a donor gain at position +1 with an intermediate confidence score of 0.27, Pangolin predicted splice loss at 3 bp, with a high score of 0.6, while the newer SAI-10k-calc tool, which interprets SpliceAI outputs to predict splicing aberration type, the size of inserted/deleted sequence, and effect on reading frame, does not provide a prediction for this variant (Canson et al. 2023). We therefore interrogated expression via RNA-seq which showed heterozygous skipping of an in-frame exon coding for the protein’s bromodomain adjacent to the intronic variant (**Fig. 3D**). We additionally leveraged the base-level methylation status information available in the HiFi LR-GS dataset to examine the CpG sites comprising the episignature of Chung-Jansen syndrome (Vos et al. 2024). This sample was a clear outlier (Z-score = 2.74) in terms of methylation status at these CpG sites (**Fig. 3E**), further corroborating the diagnosis.

##### Identification of clinically significant repeat expansions

LR-GS has a clear advantage over SR-GS for identification of repeat expansions (Chaisson et al. 2023; “Tandem Repeats in the Long-Read Sequencing Era” 2024). Examination of the *TRGT* tool output in the HiFi dataset identified two new diagnoses in the cohort.

#### Case 4 (PMGRC-1284-1284-0)

A male in their 30’s was found to harbor 460/13 CAGG repeats in *CNBP*, within the established pathogenic range (75-11,000 repeats) for myotonic dystrophy type 2 (MIM#602668) (Todd and Paulson 2010). Current phenotype includes pancreatic insufficiency, Ehlers-Danlos syndrome, aortic root dilation, executive function deficit, paresthesia, memory loss, frequent infections in childhood.

#### Case 5 (PMGRC-1067-1067-0)

In the second family, we detected 615/0 and 649/0 TAGAA repeats, in the proband and one parent respectively, in *BEAN1*, consistent with spinocerebellar ataxia 31 (MIM#117210), a condition with incomplete penetrance and late onset (Sato et al. 2009). The proband is a female in their 20’s with increased fatigue, unstable gait, use of electric wheelchair, decrease in mitochondrial complex activity, exercise intolerance and dilated cardiomyopathy. The parent is reported to be healthy at this time.

### LR-GS revealed an incidental finding not detectable with SR-GS

Cohort-wide examination of repeat expansions in the LR-GS dataset also detected a pathogenic CAG repeat expansion in *HTT* in one parent of a proband. This parent carried alleles with 20 and 41 repeats, the latter exceeding the established threshold for Huntington’s disease (≥37 repeats) (Duyao et al. 1993). This finding was unrelated to the primary indication for testing, highlighting that LR-GS may reveal clinically relevant repeat expansions that should be considered incidental findings.

### LR-GS illustrates the diagnostic utility of a single test that simultaneously interrogates genomic and DNA methylation variation

In addition to the Chung-Jansen syndrome case (Case 3, above) which was diagnosed through a combination of *duoNovo*, RNA-seq, and methylation profiling, disease-specific methylation patterns provided critical diagnostic information in three additional cases (**Fig. 4**). In **PMGRC-611-611-0**, a young female with developmental delay, hypotonia, ADHD, obsessive compulsive behaviors and history of medullary pilocytic astrocytoma, was found to have a heterozygous variant (NM_022552.5:c.1758C>G;p.Cys586Trp) in *DNMT3A* (#MIM602769) of unknown inheritance. We analyzed 139 CpG sites specifically associated with a Tatton-Brown-Rahman syndrome (MIM#615879) episignature previously reported in the literature (Husson et al. 2024). Comparing the methylation levels of these sites in the proband versus all other blood samples from the GREGoR cohort (n=162), we found that the proband was an outlier (**Fig. 4A**), consistent with a pathogenic effect of the variant. While this diagnosis could have been delivered with SR-GS followed by array-based EpiSign testing, our finding highlights the utility of multi-omic testing enabled by LR-GS, without the need for serial testing with different modalities.

**Figure 4.**
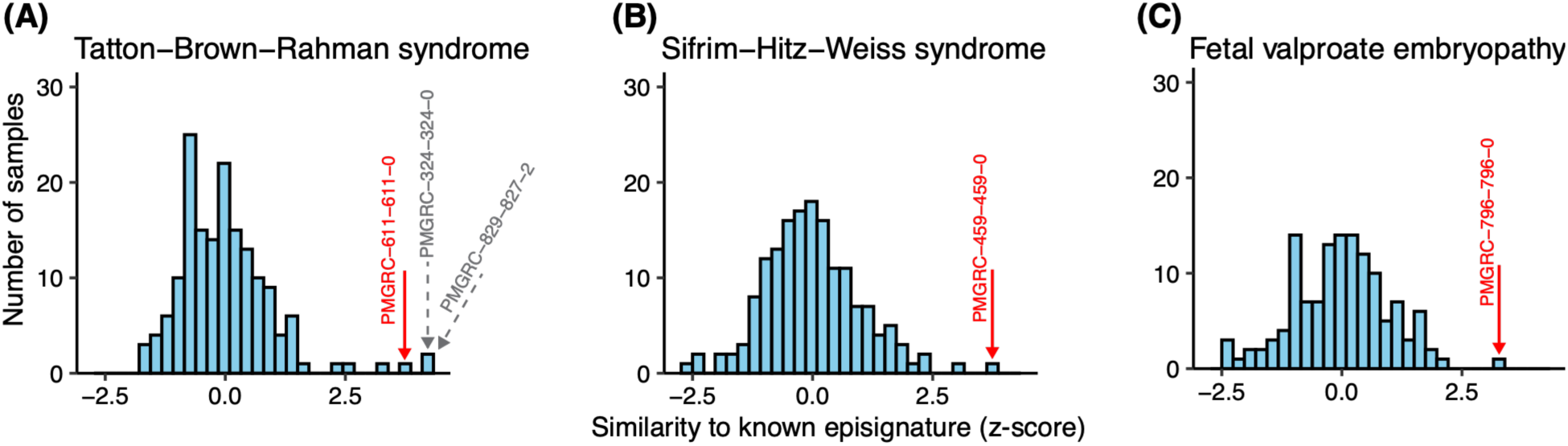
LR-GS-based methylation analysis enables the detection of probands with methylation similarity to known disease-specific episignatures. **(A) – (C)** Episignature similarity z-scores for the probands (indicated by the red arrows), against the distribution of similarity z-scores for the rest of the cohort. In panel (A), two additional samples with outlier methylation similarity to the Tatton-Brown-Rahman syndrome episignature were detected and are indicated by the gray arrows.

Another proband **(PMGRC-459-459-0)**, a young female with childhood apraxia of speech, hypertonia and autonomic neuropathy had previously been found to harbor a *de novo* heterozygous variant (NM_001273.5:c.3517C>T;p.Arg1173Trp) in *CHD4* (#MIM603277), which had been classified as a VUS. After the LR-GS-based episignature revealed an outlier methylation pattern at the relevant CpGs for Sifrim-Hitz-Weiss syndrome ((Karimi et al. 2025); **Fig. 4B**), the variant was reclassified as likely pathogenic based on the PP4 criterion (Karimi et al. 2025; Kerkhof et al. 2024; Brnich et al. 2019). While this variant thus likely explains at least part of the phenotype, the case remains unsolved because of the imperfect phenotypic match with published reports.

Finally, in PMGRC-796-796-0, a young male with autism, dysmorphic features, limb malformations including thumb hypoplasia, global developmental delay, hypotonia, nystagmus, gut malrotation and sensory disturbances, we found an outlier methylation signature (**Fig. 4C**) consistent with prenatal fetal valproic acid exposure (van der Laan et al. 2025). This prompted us to obtain further clinical history, which confirmed that the birth mother was in treatment for her bipolar disorder with valproic acid throughout the pregnancy. This highlights the potential of LR-GS to identify diagnoses for both genetic conditions and teratogenic exposures in a single assay.

### Eight cases received a diagnosis that could have been discovered with SR-GS reanalysis

In addition to diagnoses that critically relied on the unique capabilities of LR-GS, we found eight diagnoses that reanalysis of the SR-GS data could also have identified. Two cases were diagnosed because of new gene-disease associations. One was a frameshift case of *GIGYF1*-related disorder (NM_001375765.1:c.332del (p.Leu111fs)), in a male proband whose phenotype included global developmental delay, autism, seizures, dystonia, ataxia and Chiari malformation (PMGRC-559-559-0). GIGYF1-related disorder is an extremely rare condition, with under 100 cases reported globally (https://www.gigyf1.org/what-is-gigyf1) (not yet curated into OMIM; (G. Chen et al. 2022; Ding et al. 2023). The second one, which we have already reported (Boukas et al. 2026), was a case (PMGRC-730-730-0) of the recently described developmental and epileptic encephalopathy 119 (DEE119, MIM#621304), caused by a variant in *RNU2-2* (#MIM621238), which encodes the U2-2 small nuclear RNA. The variant detected was n.4G>A, one of the pathogenic variants reported in the original disease characterization study (Greene et al. 2025). During our LR-GS analysis, a genome-wide screen for putative disease-causing *de novo* variants using *duoNovo* identified this variant as *de novo*, leading to the diagnosis.

### Identification of new candidate genes and variants

We identified candidate variants in disease-associated genes and novel genes in 16 additional cases (Supp. Table 2). Of those variants, one was undetectable with SR-GS. This variant was an inversion in *trans* with a previously identified missense variant in *UBAP1* (#MIM609787), a gene that has not yet been associated with an autosomal recessive phenotype. The remaining candidate variants could all have been detected via reanalysis of SR-GS.

## Discussion

Our study highlights that the known advantages of LR–GS in the detection and interpretation of variation indeed translate into an increased diagnostic yield when directly compared with SR-GS. Specifically, LR-GS solved five previously undiagnosed cases (4.7%) by virtue of distinct capabilities: detection of complex structural variants, detection of variants in regions unmappable with SRs, detection of *de novo* variants without sequencing of both biological parents, and analysis of methylation patterns.

In these newly solved cases, patients had several prior genetic tests that failed to provide a diagnosis, including SR-GS. As a result, our 4.7% estimate represents the incremental yield of LR-GS over SR-GS, while the overall yield is substantially higher. Related to this, no previously detected diagnostic variants were missed by LR-GS. This work therefore illustrates the potential for long-read platforms to replace multiple other testing platforms, as proposed elsewhere to consolidate the information traditionally obtained from multiple genetic tests into a single assay (Damaraju et al. 2024; Abuijlan et al. 2026; Del Gobbo and Boycott 2025). Therefore, when considering the cost of diagnostic LR-GS, it should be compared to the cumulative financial and emotional cost of successive genetic tests. Notably, for the growing list of disorders associated with a diagnostic episignature, LR-GS simultaneously detects sequence and DNA methylation variation on the same DNA molecule. Analyzing these data together can accelerate the diagnostic process, using the established episignatures for several neurodevelopmental disorders to identify disorder-specific methylation patterns (Wang et al. 2026; Geysens et al. 2025).

Interestingly, two other samples (not included in the current report) also showed outlier status (**Fig. 4A**) at the sites reported in the Tatton-Brown-Rahman Syndrome episignature (Kumps et al. 2023). In one of them, we found a pathogenic *DMNT3A* variant, which had not been prioritized on prior testing; however, it is not a reasonable phenotypic match. The other was found in an unaffected unrelated mother, and thus its significance is unclear. While all early reports were of episignatures specific to the syndromes in which they were discovered and where the observed patterns were found consistently in all affected individuals there is now strong evidence that signatures of various NDD conditions are overlapping (Aref-Eshghi et al. 2021).

Episignatures have been defined to date using a microarray-based assay, which tests methylation status at 850,000 of the ∼30 million CpGs in the human genome. The increasing number of genome-wide methylomes contained in LR-GS datasets, may very well be refining these episignatures. (In this cohort, *pb-CpG-tools* reported ∼29,000,000 calls on average per genome).

The fact that eight findings identified by LR-GS that could have been solved by SR-GS echoes a common theme of genetic testing: that increased diagnostic yield is driven by advances in both detection and interpretation (Marwaha et al. 2022). LR-GS reveals large amounts of genetic sequence, but our interpretation is still limited by the phenotypic and gene-disease information available at the time of analysis. Therefore, reanalysis of existing data at a later date can reveal new diagnoses and tools are now being developed to enable automated reanalysis at scale (Welland et al. 2026; S. M. Hiatt et al. 2018; Best et al. 2024). Additionally, technological advances in variant detection can be leveraged best in the context of broader shared knowledge about population variation (e.g. gnomAD) and gene-disease relationships (e.g. GenCC/ ClinGen). Candidate variants from this study were submitted to MatchMaker (Philippakis et al. 2015) and genetic and meta-data is shared in ANViL (<u>AnVIL Portal</u>) to support ongoing re-analysis and discovery in the currently unsolved cases.

As expected, the ability of LR-GS to solve undiagnosed cases comes with an increased likelihood of incidental findings. The incidental finding of a repeat expansion pathogenic for Huntington’s disease serves as a cautionary reminder that, while LR-GS increases our ability to detect diagnostic findings, it also increases the risk of incidental findings. This highlights the importance of informed consent in the enrollment process and appropriate counseling at the time of return of results.

## Conclusions

Our study adds to the growing body of evidence indicating that a multiomics approach can be particularly powerful for diagnostic purposes, especially when combining genome sequencing, epigenomics, and RNA-seq for direct interrogation of variant effects on DNA methylation, gene expression and splicing. We envision that future developments systematizing the LR-GS-based analysis of episignatures, the analysis of RNA-seq for each sample, and incorporating Paraphase (to map variants in segmentally duplicated regions; (X. Chen et al. 2025, 2023)) and *duoNovo* (for detection of *de novo* variants when a trio is not available; (Boukas et al. 2026)), together with new tools to score the impact of non-coding variants, will all be critical to fully leverage the power of LR-GS in rare disease diagnosis.

## Declarations

### Availability of data and materials

All participants consented for general research use data sharing. The datasets supporting the conclusions of this article are available in the GREGoR workspace in AnVIL (<u>AnVIL Portal</u>). Alignments and variant call files are available on the NHGRI Analysis Visualization and Informatics Lab-space (AnVIL; <u>AnVIL Portal</u>) on the GREGoR Consortium workspace (dbGap accession <u>phs003047.v4</u>).

### Data Share Statement

Prioritized variants were discussed during a weekly meeting of a multidisciplinary team to decide on next steps, including as return of results, deposition of interpreted variants in ClinVar, or sharing of candidates in MatchMaker Exchange.

## Data Availability

All participants consented for general research use data sharing. The datasets supporting the conclusions of this article are available in the GREGoR workspace in AnVIL (AnVIL Portal). Alignments and variant call files are available on the NHGRI Analysis Visualization and Informatics Lab-space (AnVIL; AnVIL Portal) on the GREGoR Consortium workspace (dbGap accession phs003047.v4).

https://anvilproject.org/

## Acknowledgements

We thank the participants and referring physicians for helping to make this study possible.

## Funding

The study was supported by the National Institutes of Health grant U01HG011745, as part of the GREGoR Consortium. KMB was supported by the GREGoR Consortium Research Grant from the GREGoR Data Coordinating Center [U24HG011746]; National Eye Institute [R01EY035717 (KMB) and P30EY014104 (MEEI core support)], Iraty Award 2023 (KMB), Lions Foundation (KMB) and the Research to Prevent Blindness Unrestricted Grant (KMB).

### Ethics approval and consent to participate

This study was approved by the Children’s National Hospital Institutional Review Board (IRB) under protocol Pro00015852. Informed consent was obtained from all participants as required by the IRB.

### Author Contributions

GP, KB, RB, NK, SLS, AG and ES performed variant curation and interpretation. IDD, SB, SJ, JH, KC generated and processed data. IDD performed the initial variant annotation. AK and SIB processed RNA-seq data. LB and SIB performed the *duoNovo* analyses. SIB processed and analyzed methylation data. JA performed the urine amino acid analysis. KMB performed the mini-gene splicing assay. GP wrote the initial manuscript draft with input from SB and IDD, which was then revised and edited by GP, KB, RB, SB, SLS, ES, HCK, JL, LB, SIB, ED, CX. RK, SIB, ED, CX and EV supervised the study. EV obtained funding.

## Supplemental Figures and Tables

**Supplemental Figure 1.**
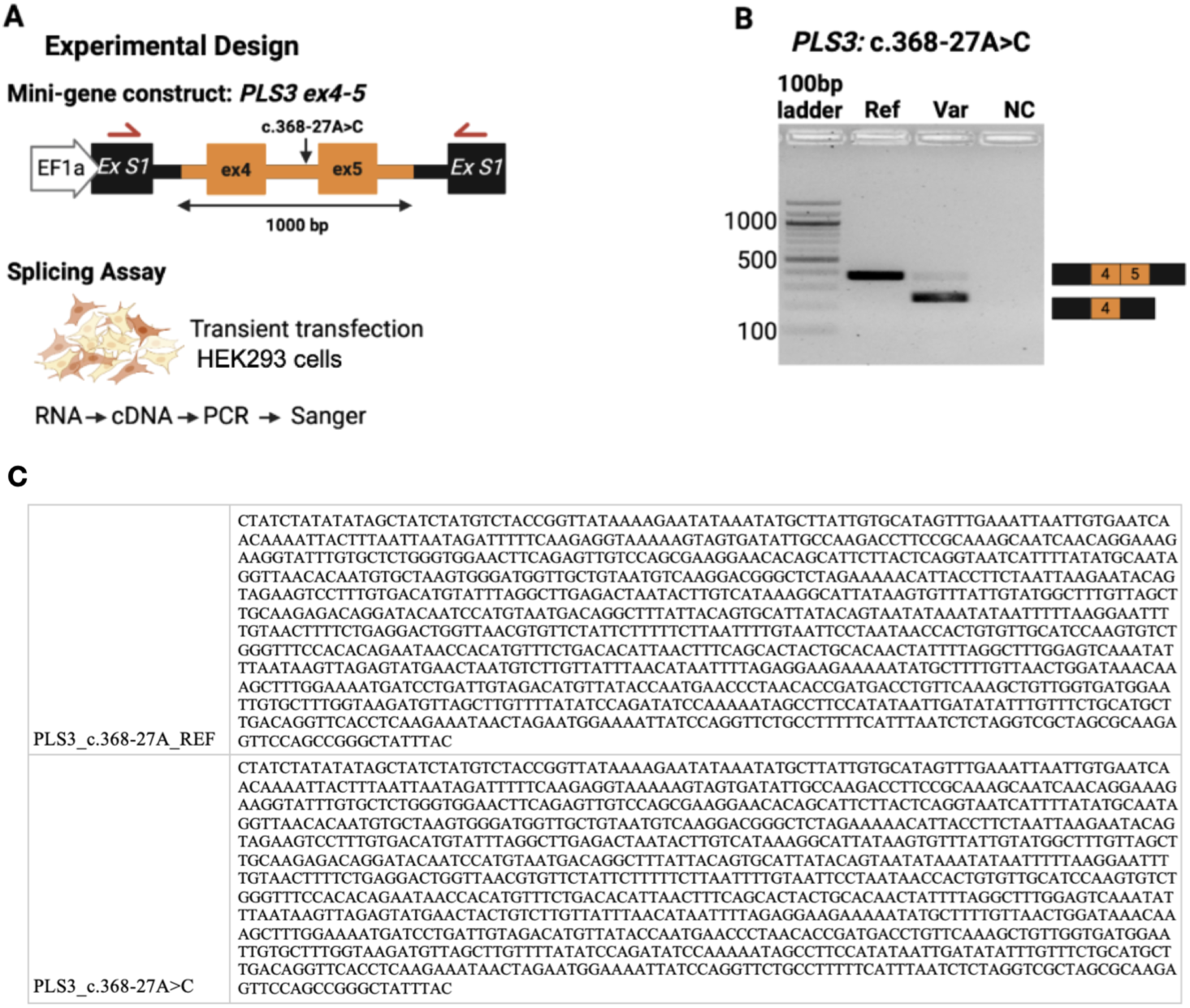
Mini-gene splicing assay to confirm impact of the intronic variant under consideration on splicing of *PLS3*. (A) The mini-gene construct used for transient transfection into HEK293 cells. Following transfection, total RNA was extracted, reverse-transcribed, and amplified to visualize splicing events on electrophoresis (Scott et al. 2022; Chong et al. 2019), in which N and C-terminal parts of the *GFP* gene were separated by *SMN1* introns 7 and 8 (NM_000344). Reference and variant (*PLS3* c.368-27A>C) gene fragments containing 1000 bp of *PLS3* exons 4 and 5 (ENST00000355899.8) and surrounding intron sequences flanked with 30-bp vector homology arms were synthesized (TWIST Bioscience, USA) and cloned into the mini-gene construct (Gibson Assembly Master Mix, New England Biolabs). After Sanger sequencing verification of all constructs, they were transfected into the HEK293 cells (Lipofectamine 3000, Thermo Fisher Scientific). Forty-eight hours post transfection, total RNA was extracted from the transfected cells (RNAeasy Mini Kit, Qiagen) and cDNA was generated using random hexamer primers (SuperScript IV Synthesis Kit, Thermo Fisher Scientific). Subsequently, the minigene transcripts were amplified from the cDNA using primers specific to the split GFP fragments (F: 5’-CACACTGGTGACAACATTTACATAC-3’; R: 5’-GAAATCGTGCTGTTTCATGTGATC-3’). The PCR products were column-purified (DNA Clean & Concentrator-5, Zymo Research) and analyzed with agarose gel electrophoresis and Sanger sequencing. (B) Gel image showing the splicing patterns generated by the reference and variant alleles. In particular, the variant allele leads to a smaller spliced product, consistent with skipping of exon 5. (C) *PLS3* gene fragments.

**Supplemental Figure 2.**
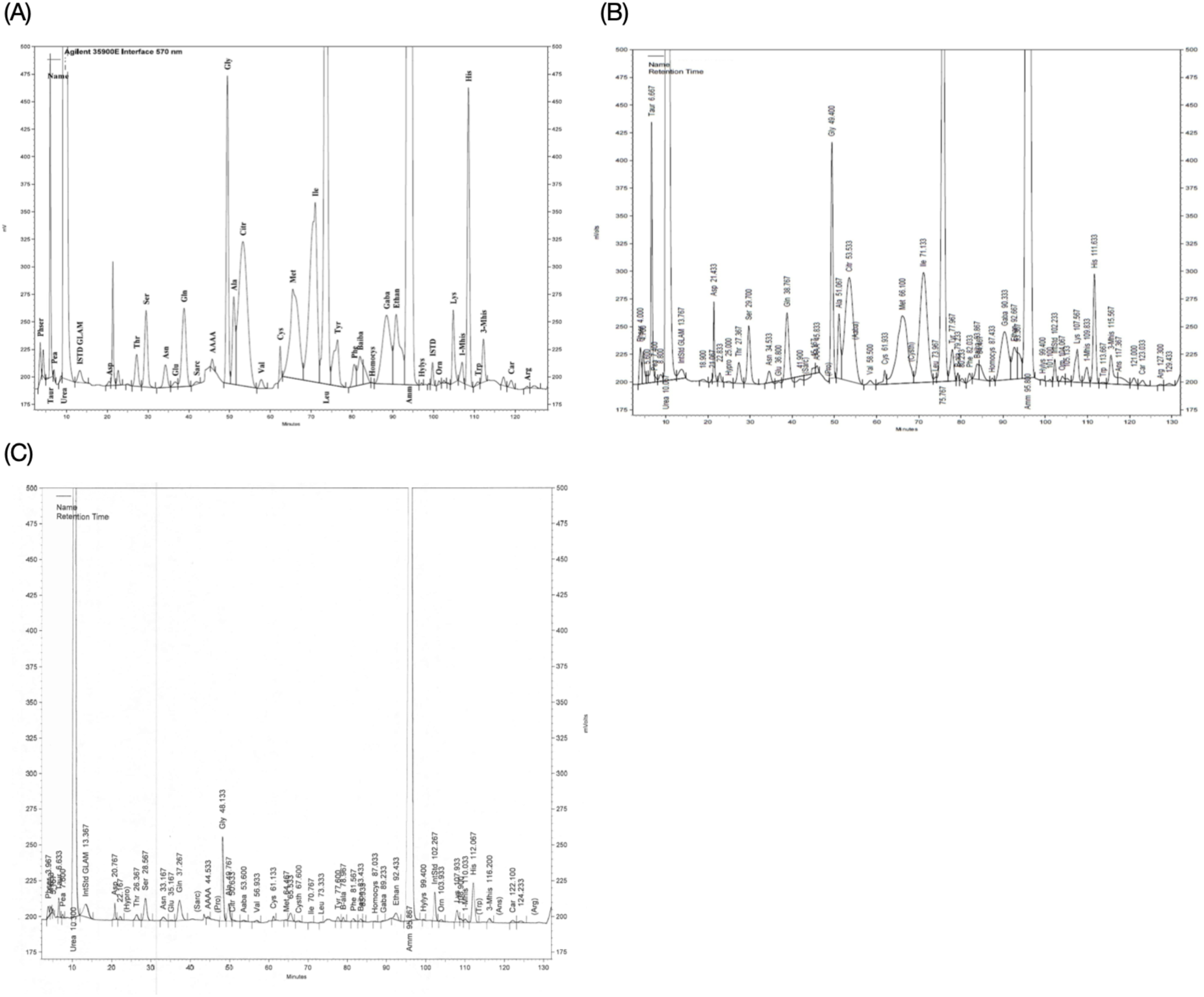
Urine amino acid analysis for biochemical interpretation of the *PEPD* variant in proband PMGRC-645-645-0. (A) Harvard Bioscience biochrom amino acid trace of urine from the proband. The presence of broad interfering peaks for many of the identified peptides indicates the presence of dipeptides, enabling the diagnosis of imidodipeptiduria. **(B)** Trace of urine from the affected sibling. Similarly to the proband, the presence of broad interfering peaks for many of the identified peptides enables the same diagnosis of imidodipeptiduria. **(C)** Calibration trace of a reference urine sample. The narrower peaks for the individual peptides identified versus those in the affected samples above can be seen.

**Supplemental Table 1.**
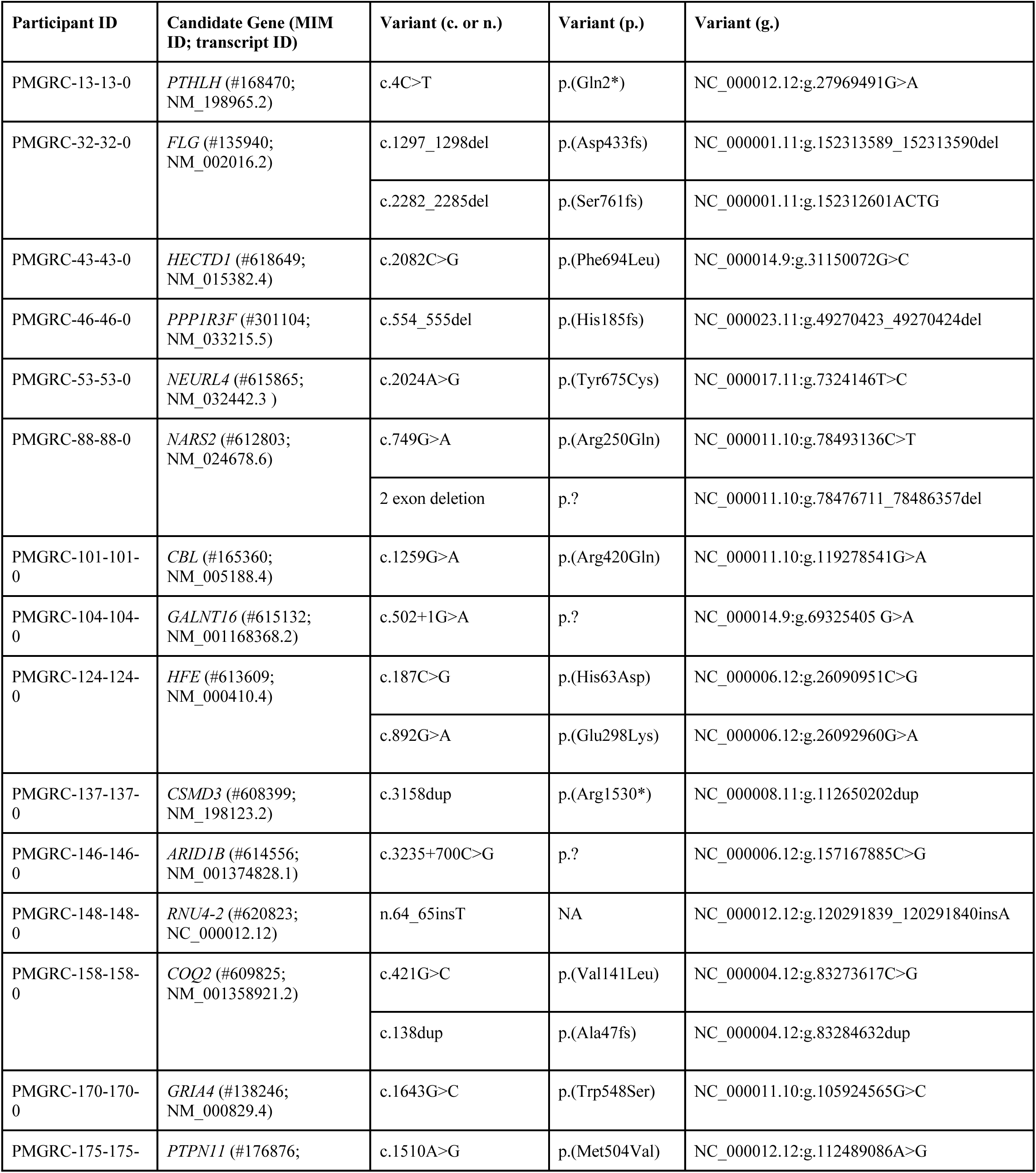
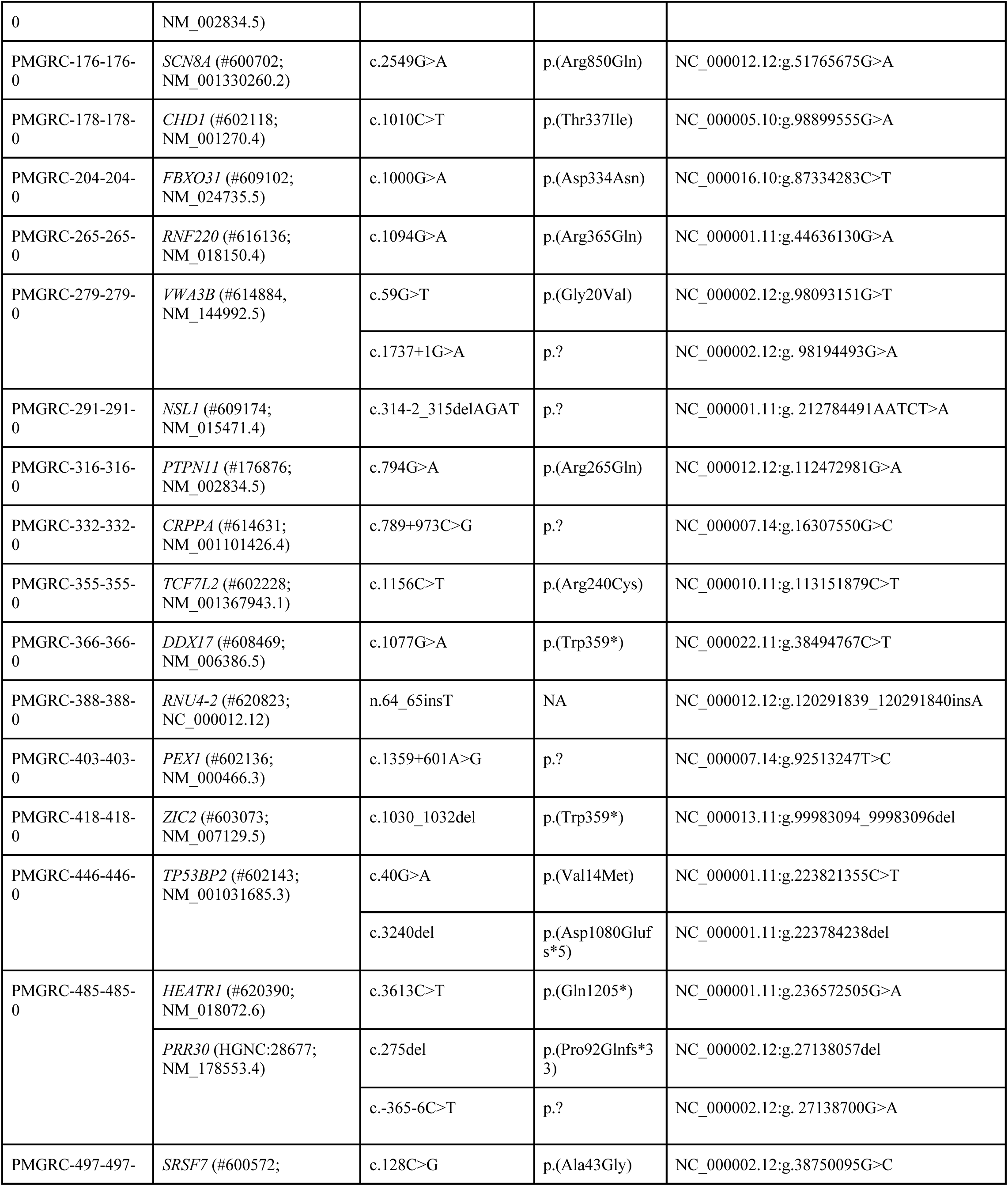
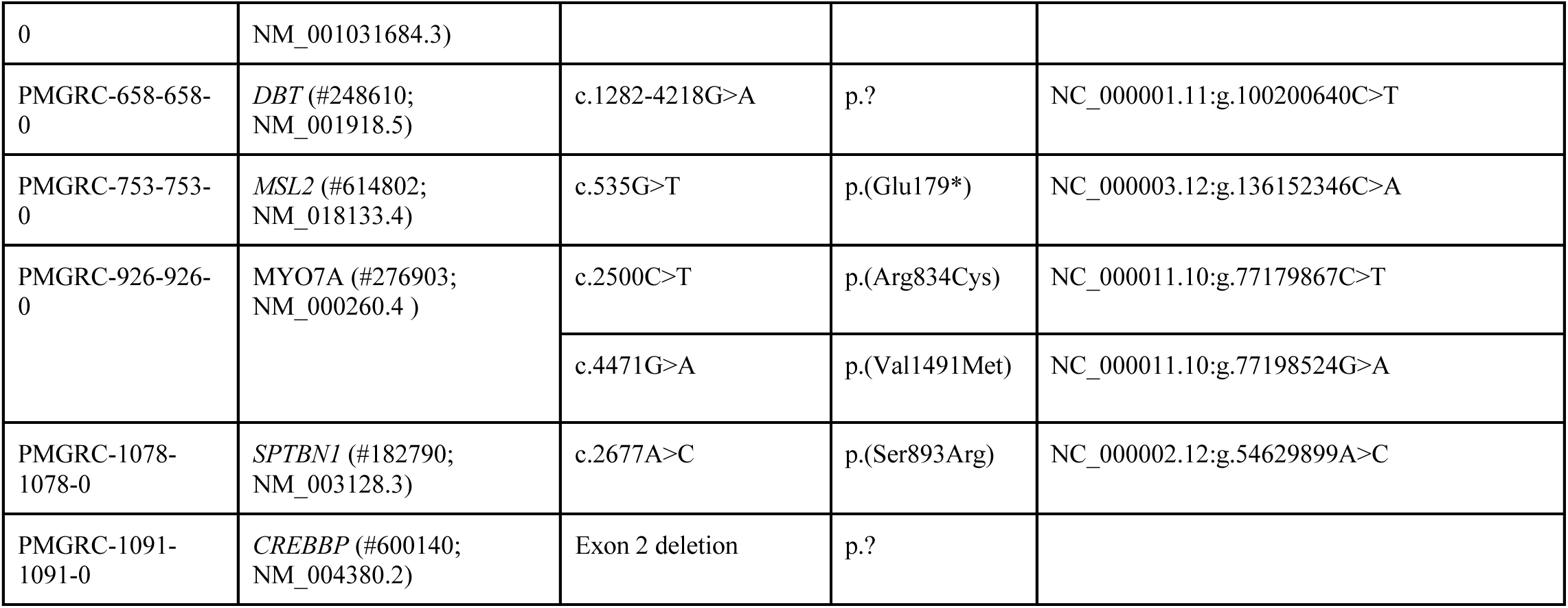
Cases with a positive finding in SR-GS.

**Supplemental Table 2.** Candidate genes.

| Participant ID | Candidate Gene (MIM ID; transcript ID) | Variant (c. or n.) | Variant (p.) | Variant (g.) | ACMG Classification |
| --- | --- | --- | --- | --- | --- |
| PMGRC-294-294-0 | DENND4B (#619843, NM_014856.3) | c.2703_2704insGCA G | p.Gln902fs*63 | NC_000001.11:g.153934829_153934830insCTGC | VUS |
| PMGRC-299-299-0 | GDNF (#600837, NM_000514.4) | c.240T>A | p.Asp80Glu | NC_000005.10:g.37816047A>T | VUS |
| PMGRC-411-411-0 | PPP1R12A (#602021, NM_002480.3) | c.737A>C | p.Glu246Ala | NC_000012.12:g.79828375T>G | VUS |
| PMGRC-429-429-0 | KIF7 (#611254, NM_198525.3) | c.2653G>A | p.Ala885Thr | NC_000015.10:g.89633206C>T | VUS |
|  |  | c.595G>A | p.Gly199Ser | NC_000001.11:g.35893756G>A | VUS |
| PMGRC-445-445-0 | ZRSR2 (#300028, NM_005089.4) | c.42-11T>A | p.? | NC_000023.11:g.15790923T>A | VUS |
| PMGRC-462-462-0 | EIF3M (#609641, NM_006360.6) | c.65T>C | p.(Leu22Pro) | NC_000011.10:g.32587034T>C | VUS |
| PMGRC-482-482-0 | GRIN2A (#138253, NM_001134407.3) | c.414+68001_414+68002insTTT | p.? | NC_000016.10:g.10111996_10111997insAAA | VUS |
| PMGRC-608-608-0 | UBAP1 (#609787, NM_016525.5) |  | p.? | NC_000009.12:g.34245081_34274111inv | VUS |
| PMGRC-641-641-0 | PHIP (#612870, NM_017934.7) | c.4631-1G>A | p.? | NC_000006.12:g.78945498C>T | VUS |
| PMGRC-674-674-0 | BAZ2B (#605683, NM_013450.4) | c.5515_5535delinsATC | p.Ser1839_Lys1845delinsIle | NC_000002.12:g.159337692_159337712delinsGAT | VUS |
| PMGRC-772-772-0 | C1QBP (#601269, NM_001212.4) | c.829_830del | p.(Ser277PhefsTer23) | NC_000017.11:g.5433036_5433037del | VUS |
| PMGRC-773-773-0 | ZNF609 (#617474, NM_015042.2) | c.1096C>A | p.(Arg366Ser) | NC_000015.10:g.64673950C>A | VUS |
| PMGRC-861-861-0 | MEF2A (#600660, NM_001319206.4) | c.764C>G | p.(Pro255Arg) | NC_000015.10:g.99690334C>G | VUS |
| PMGRC-1085-1085-0 | SSBP3 (#607390, NM_145716.4) | c.927+1G>A | p.? | NC_000001.11:g.54239128C>T | VUS |

## List of abbreviations

ACMG: American College of Medical Genetics and Genomics
AMP: Association for Molecular Pathology
AnVIL: Genomic Data Science Analysis, Visualization, and Informatics Lab-space
API: Application programming interface
AWS: Amazon Web Services
CADD: Combined Annotation Dependent Depletion
ClinGen: Clinical Genome Resource
CMA: Chromosomal microarray
CNV: Copy-number variant
GenCC: Gene Curation Coalition
GFP: Green fluorescent protein
HMW: High molecular weight
HPO: Human Phenotype Ontology
IRB: Institutional Review Board
JSON: JavaScript Object Notation
LR-GS: Long-read genome sequencing
SMRT: Single-molecule real-time
SNV: Single-nucleotide variant
SR-GS: Short-read genome sequencing
SV: Structural variant
TRGT: Tandem Repeat Genotyping Tool
VCF: Variant Call Format
VUS: Variant of uncertain significance
WDL: Workflow Description Language

